# Exploring the Relationship Between Smoking and Poor Sleep Quality: A Cross-Sectional Study Using NHANES

**DOI:** 10.1101/2024.03.17.24304413

**Authors:** Haoxiong Sun

## Abstract

**Introduction:** Although some previous research has reported the relationship between smoking and sleep disorders, there is a lack of studies with large sample sizes and determining the potential dose-response relationship.

**Methods:** This study used data from 30,269 participants from the NHANES database (2007-2020). Logistic regression models were used to assess the associations between smoking and sleep outcomes, including insufficient sleep duration, sleep problems, snoring, and snorting or stop breathing during sleep. Dose-response relationships were explored using restricted cubic splines.

**Results:** Light-to-moderate and heavy smokers had significantly higher odds of experiencing sleep disorders compared to non-smokers. The odds ratios for Light-to-moderate and heavy smokers are 1.341 and 2.101 for insufficient sleep duration, 1.493 and 2.142 for sleep problems, 1.444 and 2.003 for snoring, and 1.439 and 1.720 for snorting or stop breathing during sleep, respectively. Dose-response analyses revealed that the odds of these sleep outcomes increased with higher smoking frequency.

**Conclusion:** Smoking can lead to increased odds of sleep disorders, and the more an individual smokes, the higher the odds of experiencing sleep disorders.

## Introduction

Sleep is a vital physiological process that plays a crucial role in maintaining physical and mental well-being (Baranwal, Yu, & Siegel, 2023; Worley, 2018). Poor sleep quality has been associated with various adverse health outcomes, including cardiovascular diseases (Cappuccio et al., 2011), metabolic disorders like diabetes (Knutson & Van Cauter, 2008), and cognitive impairments (Yaffe, Falvey, & Hoang, 2014). In recent years, there has been growing interest in identifying modifiable risk factors that contribute to poor sleep quality (Itani et al., 2017).

Smoking, a prevalent global health issue, has been linked to numerous chronic diseases and premature deaths (West, 2017). Despite the well-established harmful effects of smoking on several diseases including respiratory and cardiovascular health (Parmar et al., 2023), its relationship with sleep quality remains understudied. Previous research has suggested that smoking may disrupt sleep architecture and lead to sleep disturbances (McNamara et al., 2014; Zhang et al., 2006). Understanding the relationship between smoking and sleep quality is crucial for developing targeted interventions and public health strategies to improve sleep health and overall well-being. Although some studies have identified pathological and epidemiological links between smoking and sleep disorders, there is still a lack of large-sample observational studies to comprehensively explore this association (Dugas et al., 2017).

The National Health and Nutrition Examination Survey (NHANES), a nationally representative survey conducted in the United States, collects comprehensive data on various health-related factors, including smoking habits and sleep parameters (Wu et al., 2023). By leveraging NHANES data from 2007-2020 March, we can explore the potential link between smoking and poor sleep quality while accounting for potential confounders such as age, gender, race/ethnicity, education level, marital status, and body mass index (BMI) (You et al., 2023).

In this study, we aim to investigate the association between smoking and poor sleep quality or sleeping disorders using data from multiple NHANES survey cycles. We hypothesize that individuals who smoke will have a higher prevalence of poor sleep quality compared to non-smokers.

## Method

### 2.1 Study subjects

This cross-sectional study used data from the National Health and Nutrition Examination Survey (NHANES), a nationally representative survey conducted by the Centers for Disease Control and Prevention (CDC) in the United States. NHANES employs a complex, multistage probability sampling design to assess the health and nutritional status of the civilian, non-institutionalized U.S. population (CDC, 2021).

Data from six consecutive NHANES cycles (2007-2008, 2009-2010, 2011-2012, 2013-2014, 2015-2016, and 2017-2020 March) were combined for this study, yielding a total sample size of 66,148 participants. Among these, 30,269 subjects had complete data on smoking status (including non-smokers) and were included in the analysis. Subjects who did not provide information on their smoking status (n = 35,879) were excluded. The study population varied depending on the sleep outcome of interest. For the analysis of sleep duration, 22,745 subjects with valid sleep duration data were included. The analysis of sleep problems included 22,796 subjects with available data. For the analyses of snoring and snorting or stop breathing during sleep, 8,357 and 8,468 subjects were included, respectively, based on the availability of relevant data.

All participants provided written informed consent, and the NHANES study protocol was approved by the National Center for Health Statistics Research Ethics Review Board (CDC, 2024).

### 2.2 Exposure Measurement

Smoking status was determined using the NHANES questionnaire item "SMQ020 - Smoked at least 100 cigarettes in life". Participants who reported having smoked less than 100 cigarettes in their lifetime were classified as "non-smokers". Among those who had smoked at least 100 cigarettes, the intensity of smoking was assessed using the questionnaire item "SMD650 - Avg # cigarettes/day during past 30 days".

Based on the responses to SMD650, smokers were further categorized into two groups: light-to-moderate smokers, who reported smoking more than 0 but fewer than 10 cigarettes per day, and heavy smokers, who reported smoking 10 or more cigarettes per day. These categories were used in the logistic regression analyses to examine the association between smoking intensity and sleep outcomes.

For the dose-response analysis, the continuous variable derived from "SMD650 - Avg # cigarettes/day during past 30 days" was used to represent the average number of cigarettes smoked per day. This approach allowed for a more granular examination of the relationship between smoking quantity and sleep outcomes.

### 2.3 Outcome Measurement

This study examined several sleep-related outcomes, including sleep duration sufficiency, diagnosed sleep problems, and the frequency of snoring or snorting and stop breathing during sleep.

Sleep duration was assessed using two different questionnaire items across the NHANES cycles. In the 2007-2014 cycles, sleep duration was measured using the item "SLD010H - How much sleep do you get (hours)?". For the 2015-2020 cycles, sleep duration was assessed using the item "SLD012 - Sleep hours". Based on previous research, sleep duration was dichotomized into sufficient (>7 hours per night) and insufficient (≤7 hours per night) sleep (You et al., 2023). Diagnosed sleep problems were determined using the questionnaire item "SLQ050 - Ever told doctor had trouble sleeping?". Participants who reported being told by a doctor that they had trouble sleeping were considered to have a diagnosed sleep problem.

Snoring frequency was assessed using the questionnaire item "SLQ030 - How often do you snore?". Responses were categorized as never, rarely (1-2 nights per week), occasionally (3-4 nights per week), and frequently (5 or more nights per week).

Similarly, the frequency of snorting or stop breathing during sleep was measured using the questionnaire item "SLQ040 - How often do you snort or stop breathing?". Responses were categorized as never, rarely (1-2 nights per week), occasionally (3-4 nights per week), and frequently (5 or more nights per week).

### 2.4 Covariable

Covariables were chosen based on previous research (Wu et al., 2023; You et al., 2023). Demographic characteristics were extracted from the demographic questionnaire, including age, gender, race/ethnicity, marital status, family poverty income ratio, and education level. These variables were categorized as follows: Race/ethnicity: Non-Hispanic White, Non-Hispanic Black, Mexican American, Other Hispanic, and Other Race - Including Multi-Racial. Marital status: Married/living with partner, Widowed/divorced/separated, and Never married. Family poverty income ratio: Low income (<1), Middle income (1-3), and High income (≥3). Education level: Below high school, High school/GED, and College or above. Body mass index (BMI) was extracted from “BMXBMI - Body Mass Index (kg/m**2)”. BMI was treated as a continuous variable in the analysis.

### 2.5 Statistical analysis

According to the NHANES protocol, all the data were integrated into a single dataset, and data analysis considered the complex survey design and applied the suggested weighting methodology. For weighting purposes, a new variable, WTMEC, was created by combining the WTMEC2YR (if available) or WTMECPRP variables from each NHANES cycle. Observations with missing values for WTMEC, SDMVPSU (sampling units), or SDMVSTRA (strata variables) were excluded from the analysis. The svydesign() function from the survey package was used to create weighted survey design objects, specifying the sampling weights (WTMEC), sampling units (SDMVPSU), and strata (SDMVSTRA). These design objects were then used in subsequent weighted analyses to ensure that the results were representative of the U.S. population.

Odds ratios (ORs) and 95% confidence intervals (CIs) were estimated using weighted logistic regression models to assess the associations between smoking categories and sleep outcomes (insufficient sleep duration and sleep problems). Three models were constructed: a crude model (unadjusted), Model 1 (adjusted for age, gender, and race), and Model 2 (further adjusted for BMI, education level, marital status, and poverty level). The svyglm() function was used to fit these models, accounting for the complex survey design.

The dose-response relationship between smoking amount and sleep outcomes was investigated using restricted cubic splines (RCS) in weighted logistic regression models.

The lrm() function from the rms package was used to fit these models, with smoking amount treated as an RCS term and adjusting for other covariates. The Predict() function was then used to estimate the ORs and 95% CIs for sleep outcomes at different levels of smoking amount. These results were visualized using the ggplot2 package.

Ordered logistic regression models were fitted using the polr() function to evaluate the associations between smoking categories and ordinal sleep outcomes, such as snoring frequency and sleep breathing pauses. These models were adjusted for the same set of covariates as in the logistic regression models.

All statistical analyses were performed using R software version 4.3.2 (http://www.R-project.org, The R Foundation), with a significance level of P < 0.05. The survey package was used extensively to account for the complex survey design of NHANES, ensuring that the results were properly weighted and representative of the U.S. population.

## Results

### 3.1 Smoking and Insufficient sleep duration

Table 1 assesses associations between smoking levels and insufficient sleep duration, suggested that smoking status was a significant predictor across all models. The crude model revealed that light-to-moderate smokers had 1.516 times the odds (95% CI: 1.367-1.680, P<0.001) and heavy smokers had 1.974 times the odds (95% CI: 1.757-2.217, P<0.001) of experiencing insufficient sleep compared to non-smokers. Adjustments for age, gender, and race in Model 1 slightly reduced the odds for light-to-moderate smokers to 1.433 (95% CI: 1.295-1.584, P<0.001) and increased the odds for heavy smokers to 2.101 (95% CI: 1.863-2.369, P<0.001). Further adjustment for BMI, education level, marital status, and poverty level in Model 2 resulted in odds of 1.341 (95% CI: 1.205-1.493, P<0.001) for light-to-moderate smokers and 2.000 (95% CI: 1.763-2.268, P<0.001) for heavy smokers.

**Table 1.**
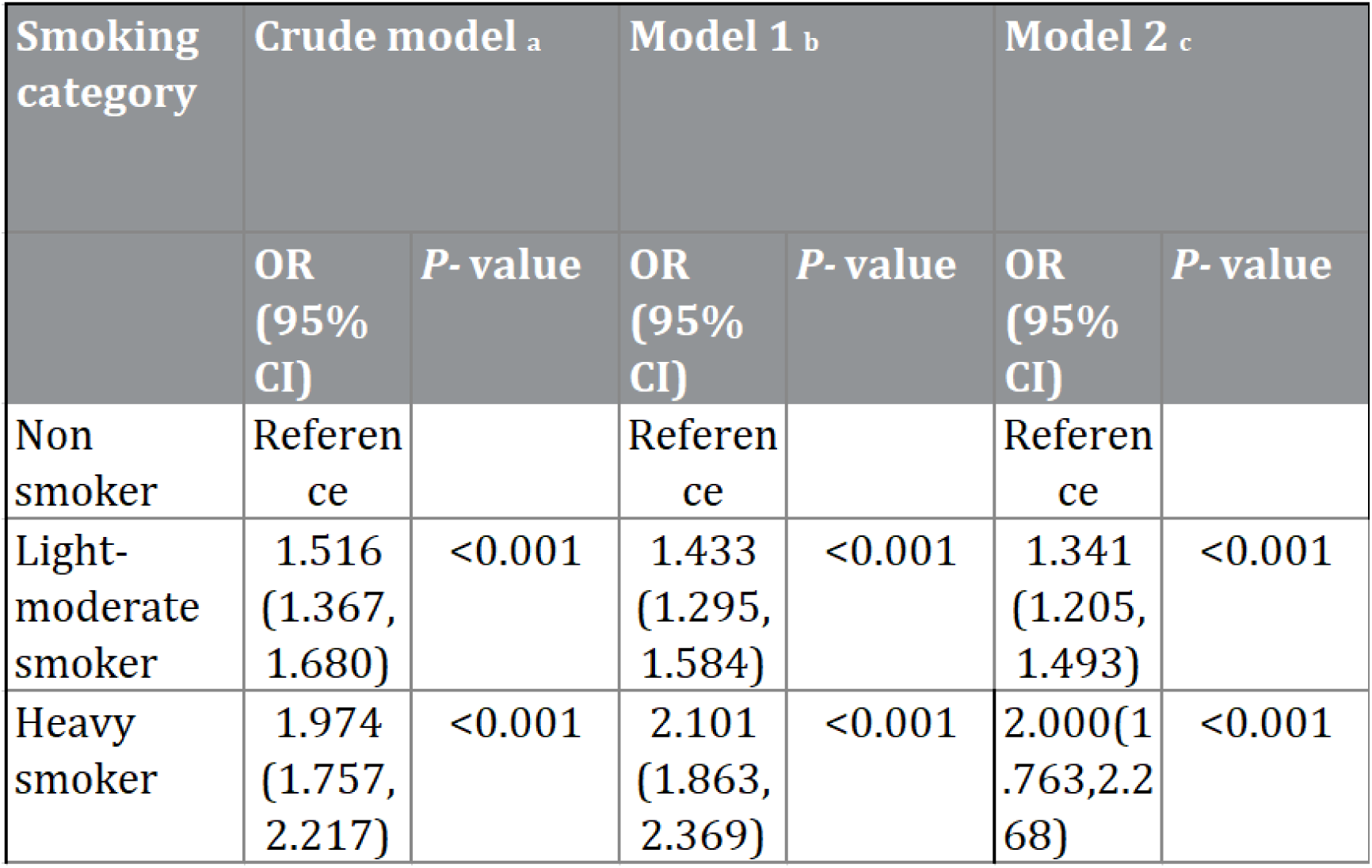
Associations between smoking levels and insufficient sleeping duration. Crude model: no covariates considered; Model 1: age, gender and race; Model 2: Age, Gender, Race, BMI, Education Level, Marital Status, Poverty Level

| Smoking category | Crude model <sup>a</sup> |  | Model 1 <sup>b</sup> |  | Model 2 <sup>c</sup> |  |
| --- | --- | --- | --- | --- | --- | --- |
|  | OR (95% CI) | P- value | OR (95% CI) | P- value | OR (95% CI) | P- value |
| Non smoker | Reference |  | Reference |  | Reference |  |
| Light-moderate smoker | 1.516 (1.367, 1.680) | <0.001 | 1.433 (1.295, 1.584) | <0.001 | 1.341 (1.205, 1.493) | <0.001 |
| Heavy smoker | 1.974 (1.757, 2.217) | <0.001 | 2.101 (1.863, 2.369) | <0.001 | 2.000 (1.763, 2.268) | <0.001 |

Figure 1 presents a dose-response curve that shows an increase in the adjusted odds ratio for insufficient sleep duration as the amount of smoking rises. This trend indicates that a higher quantity of smoking is associated with an increased probability of experiencing insufficient sleep. The expanding confidence interval suggests greater variability in the odds ratio estimates at higher smoking levels, but the overall pattern remains consistent.

**Figure 1.**
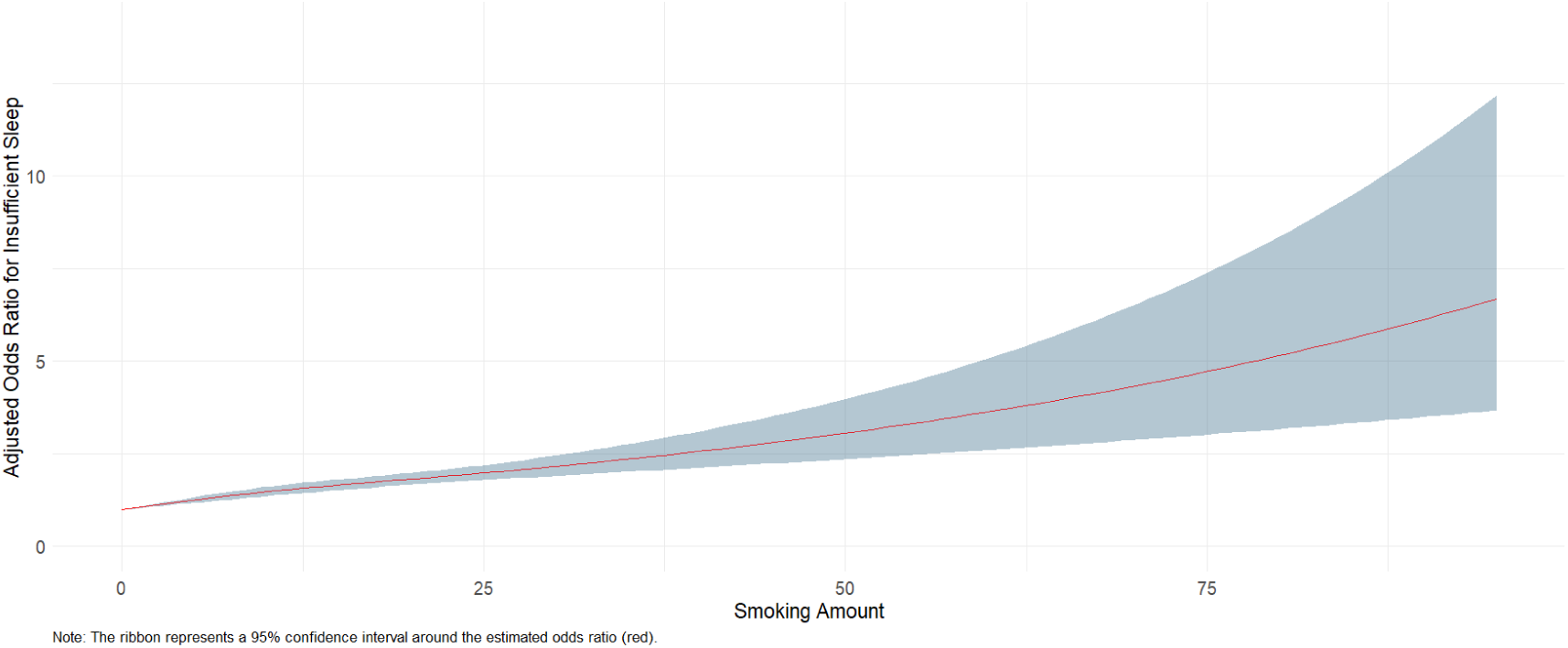
Dose-response relationship between smoking amount and sleep insufficiency.

### 3.2 Smoking and Diagnosed sleep problems

Table 2 explores the association between smoking levels and being diagnosed with sleep problems. In all models, smoking status was significantly associated with sleep problems. The crude model displayed that light-to-moderate smokers had a 49.3% increase in odds (95% CI: 1.334-1.670, P<0.001), and heavy smokers had a 97.9% increase (95% CI: 1.764-2.220, P<0.001) of having diagnosed sleep problems compared to non-smokers. Adjusting for age, gender, and race in Model 1 increased the odds for light-to-moderate smokers to 1.727 (95% CI: 1.539-1.938, P<0.001) and for heavy smokers to 2.017 (95% CI: 1.798-2.263, P<0.001). Model 2, which further accounted for BMI, education level, marital status, and poverty level, suggested that light-to-moderate smokers had 78.8% higher odds (95% CI: 1.582-2.021, P<0.001), and heavy smokers had 104.2% higher odds (95% CI: 1.805-2.310, P<0.001) of reporting sleep problems.

**Table 2.**
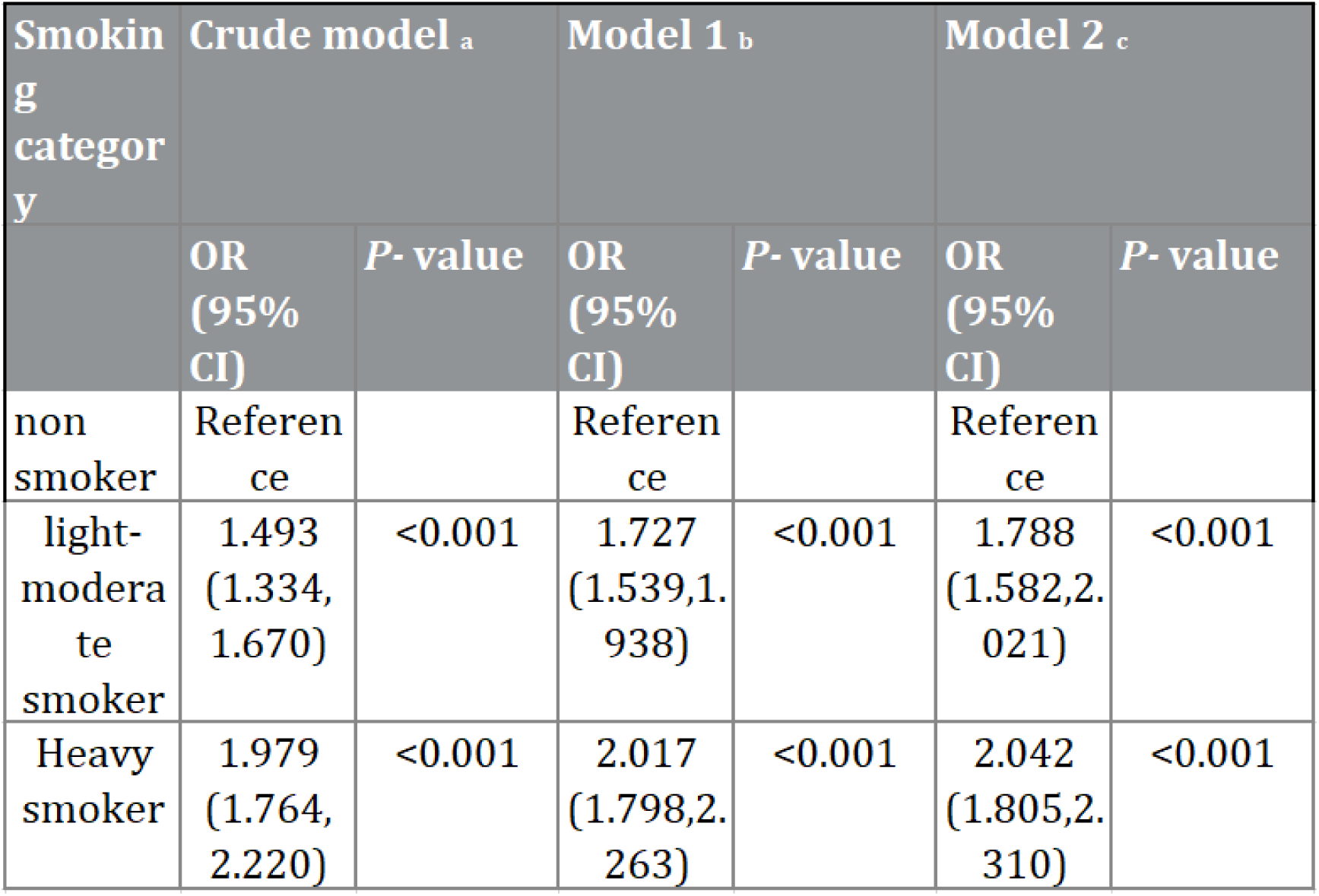
Associations between smoking levels and diagnosed sleep problems. Crude model: no covariates considered; Model 1: age, gender and race; Model 2: Age, Gender, Race, BMI, Education Level, Marital Status, Poverty Level

Figure 2 presents a dose-response curve that shows an increase in the adjusted odds ratio for the possibility of being diagnosed with sleeping problem as the amount of smoking rises. This trend indicates that a higher quantity of smoking is associated with an increased probability of experiencing sleeping problems. There is greater variability in the odds ratio estimates at higher smoking levels, but the overall pattern of increased risk with more smoking remains consistent.

**Figure 2.**
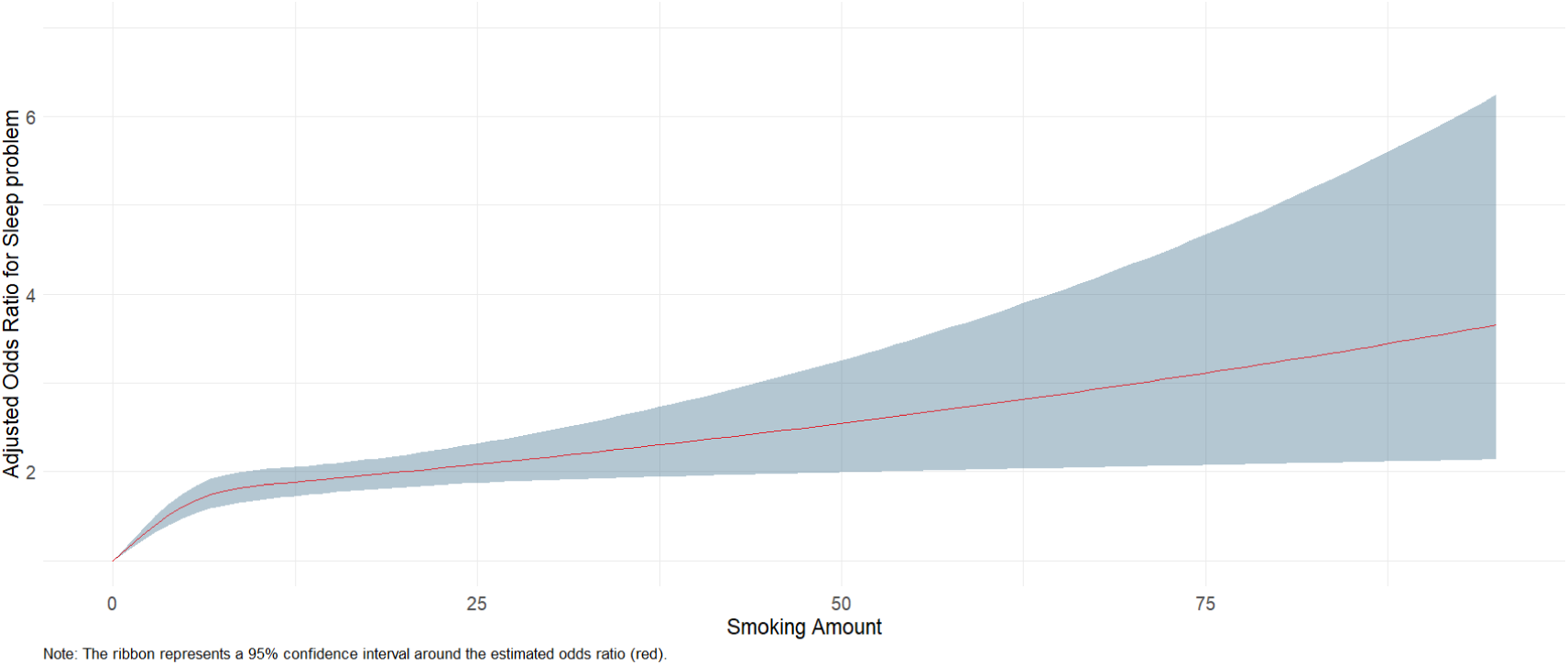
Dose-response relationship between smoking amount and diagnosed sleep problems.

### 3.3 Smoking and Snoring

Table 3 illustrates that smoking was positively linked to snoring frequency. Light-to-moderate and heavy smokers had increased odds of snoring, 1.444 and 2.003 times respectively, compared to non-smokers. Age correlated with a marginal rise in snoring odds annually (OR=1.014). Females were less likely to snore than males (OR=0.584), and variations in snoring risk were evident across races and with higher BMI, and education, marital status, and poverty levels also exhibited associations with snoring frequency.

**Table 3.**
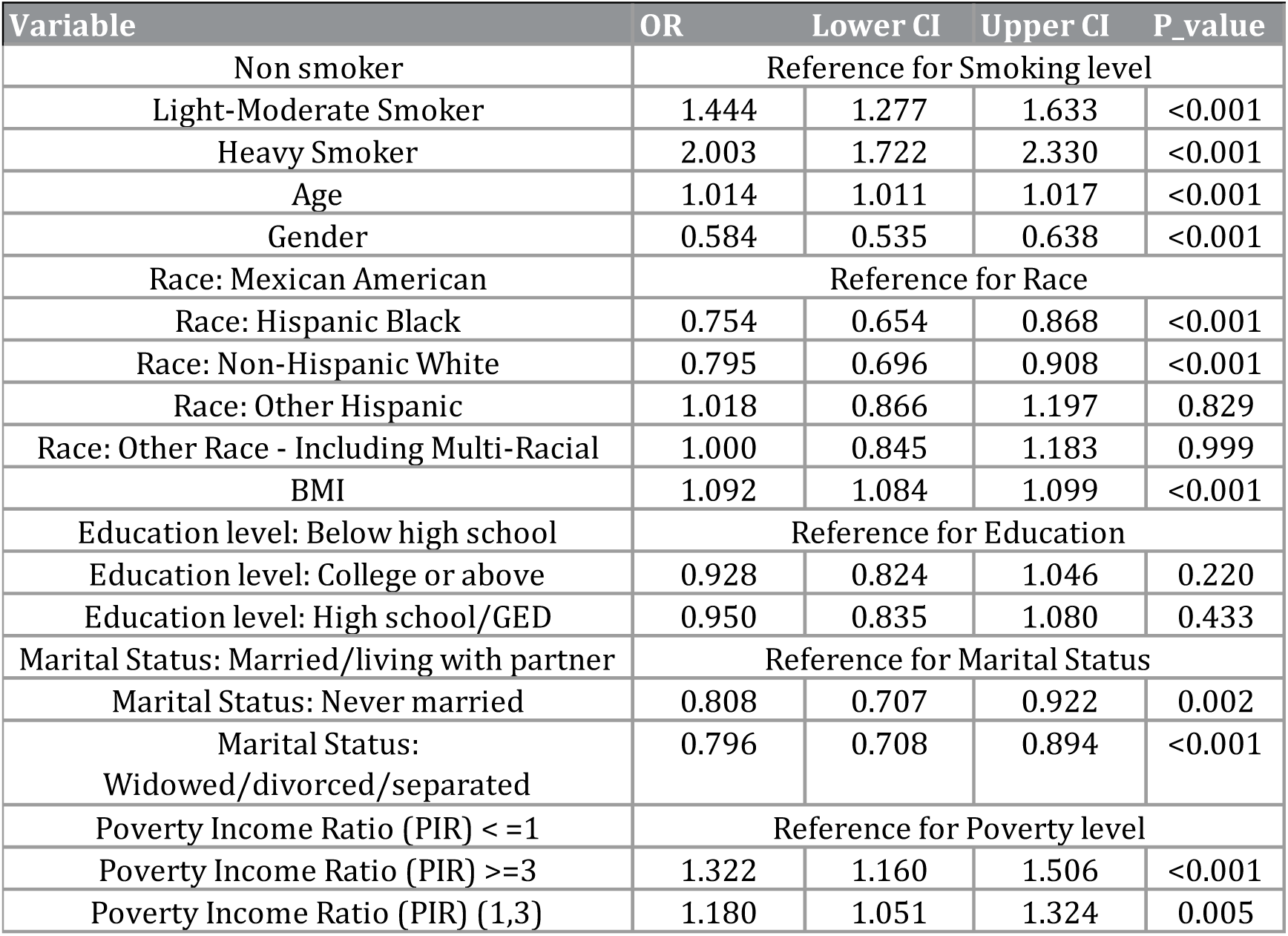
Associations between smoking levels and snoring frequency.

| Variable | OR | Lower CI | Upper CI | P_value |
| --- | --- | --- | --- | --- |
| Non smoker | Reference for Smoking level |  |  |  |
| Light-Moderate Smoker | 1.444 | 1.277 | 1.633 | <0.001 |
| Heavy Smoker | 2.003 | 1.722 | 2.330 | <0.001 |
| Age | 1.014 | 1.011 | 1.017 | <0.001 |
| Gender | 0.584 | 0.535 | 0.638 | <0.001 |
| Race: Mexican American | Reference for Race |  |  |  |
| Race: Hispanic Black | 0.754 | 0.654 | 0.868 | <0.001 |
| Race: Non-Hispanic White | 0.795 | 0.696 | 0.908 | <0.001 |
| Race: Other Hispanic | 1.018 | 0.866 | 1.197 | 0.829 |
| Race: Other Race - Including Multi-Racial | 1.000 | 0.845 | 1.183 | 0.999 |
| BMI | 1.092 | 1.084 | 1.099 | <0.001 |
| Education level: Below high school | Reference for Education |  |  |  |
| Education level: College or above | 0.928 | 0.824 | 1.046 | 0.220 |
| Education level: High school/GED | 0.950 | 0.835 | 1.080 | 0.433 |
| Marital Status: Married/living with partner | Reference for Marital Status |  |  |  |
| Marital Status: Never married | 0.808 | 0.707 | 0.922 | 0.002 |
| Marital Status: Widowed/divorced/separated | 0.796 | 0.708 | 0.894 | <0.001 |
| Poverty Income Ratio (PIR) <=1 | Reference for Poverty level |  |  |  |
| Poverty Income Ratio (PIR) >=3 | 1.322 | 1.160 | 1.506 | <0.001 |
| Poverty Income Ratio (PIR) (1,3) | 1.180 | 1.051 | 1.324 | 0.005 |

### 3.4 Smoking and Snort or stop breathing

Table 4 illustrates that the smoking levels were significantly associated with the frequency of snorting and stopping breathing during sleep. Light-to-moderate smokers were 1.439 times more likely to experience these events compared to non-smokers, while heavy smokers had 1.720 times higher. Age showed a slight but significant association with these breathing disturbances (OR=1.011), and females had a substantially lower risk than males (OR=0.584). Certain racial and ethnic groups, particularly Other Hispanics and Multi-Racial individuals, were more likely to report these events. A higher BMI was also a significant predictor (OR=1.066). Marital status showed that single individuals had lower odds compared to those married or living with a partner. Education and poverty levels did not show a significant association with the occurrence of snorting and stopping breathing during sleep.

**Table 4.** Associations between smoking levels and snoring frequency.

| Variable | OR | Lower CI | Upper CI | P_value |
| --- | --- | --- | --- | --- |
| Non smoker | Reference for Smoking level |  |  |  |
| Light-Moderate Smoker | 1.439 | 1.233 | 1.680 | <0.001 |
| Heavy Smoker | 1.720 | 1.436 | 2.060 | <0.001 |
| Age | 1.011 | 1.008 | 1.015 | <0.001 |
| Gender | 0.584 | 0.521 | 0.656 | <0.001 |
| Race: Mexican American | Reference for Race |  |  |  |
| Race: Hispanic Black | 0.848 | 0.702 | 1.024 | 0.087 |
| Race: Non-Hispanic White | 1.052 | 0.883 | 1.253 | 0.570 |
| Race: Other Hispanic | 1.312 | 1.070 | 1.608 | 0.009 |
| Race: Other Race - Including Multi-Racial | 1.315 | 1.053 | 1.642 | 0.016 |
| BMI | 1.066 | 1.057 | 1.074 | <0.001 |
| Education level: Below high school | Reference for Education |  |  |  |
| Education level: College or above | 0.984 | 0.844 | 1.147 | 0.835 |
| Education level: High school/GED | 0.963 | 0.816 | 1.136 | 0.651 |
| Marital Status: Married/living with partner | Reference for Marital Status |  |  |  |
| Marital Status: Never married | 0.743 | 0.618 | 0.892 | 0.001 |
| Marital Status:<br>Widowed/divorced/separated | 0.824 | 0.710 | 0.957 | 0.011 |
| Poverty Income Ratio (PIR) < =1 | Reference for Poverty level |  |  |  |
| Poverty Income Ratio (PIR) >=3 | 1.023 | 0.864 | 1.212 | 0.789 |
| Poverty Income Ratio (PIR) (1,3) | 0.944 | 0.814 | 1.094 | 0.443 |

## Discussion

This large, nationally representative cross-sectional study found significant associations between smoking and various sleep outcomes, including insufficient sleep duration, sleep problems, snoring, and snorting or stop breathing during sleep. Dose-response analyses revealed that the odds of these sleep outcomes increased with higher smoking frequency, consistent with previous studies that smokers would have an elevated prevalence of sleeping disorders (Cohrs et al., 2014; Jang et al., 2023; Liao et al., 2019). However, our study provides more solid evidence due to the large sample size and examination of dose-response relationships.

Several potential mechanisms may underlie the association between smoking and sleep disturbances. Nicotine, the main stimulant present within the cigarette, can increase arousal and lead to withdrawal symptoms during the night (Hughes, 2007; Jaehne et al., 2015). Smoking also affects circadian rhythms, with smokers typically having lower melatonin secretion levels, which may disrupt the sleep-wake cycle (Ursing et al., 2005). Nicotine can also interfere with acetylcholine pathway, which further disrupt the sleep-wake cycles (Salin-Pascual et al., 1999). Smoking can also activate inflammatory pathways in the body, leading to a state of chronic systemic inflammation associated with sleep disorders (Dzierzewski et al., 2020; Lee, Taneja, & Vassallo, 2012). Furthermore, smoking is associated with various mental health problems, such as anxiety and depression, which are risk factors for sleep disorders (Fluharty et al., 2017). Chronic diseases caused by smoking, such as chronic obstructive pulmonary disease (COPD) and cardiovascular disease, may also affect sleep quality through multiple mechanisms (Badran et al., 2015; Budhiraja, Siddiqi, & Quan, 2015). Lastly, smoking behavior itself may affect sleep hygiene and sleep habits, as many smokers smoke before bed or when they wake up during the night, which can delay sleep onset or disrupt sleep (Peters et al., 2011).

The dose-response relationship indicates that even if complete cessation of smoking is not achievable, reducing the amount of smoking can still positively affect sleep quality and reduce the risk of sleep disorders. Previous studies believe that the cessation rather than reduction of smoking can improve the health status, like cardiovascular status (Godtfredsen et al., 2002; Jeong et al., 2021), however, our research indicates that at least for sleep disorders, reduction in smoking amount can also improve the consequences, which provides a basis for formulating more flexible and realistic public health strategies.

Our study has several strengths, including the use of a nationally representative sample, examination of multiple sleep outcomes, and exploration of dose-response relationships. Our findings have important implications for public health and clinical practice, addressing smoking could be a key strategy for improving sleep health at the population level. However, this research contains several limitations. One limitation of this study is its cross-sectional design, which precludes the establishment of causal relationships between smoking and sleep outcomes. Additionally, the use of self-reported smoking status may introduce potential biases, such as recall bias or social desirability bias, which could affect the accuracy of the data. Finally, although we adjusted for several important confounders, the possibility of residual confounding by unmeasured factors cannot be entirely ruled out.

In conclusion, this study found significant associations between smoking and various sleep problems, with dose-response relationships. These findings underscore the importance of addressing smoking to improve sleep health and reduce the risk of related health problems. Healthcare providers should routinely assess smoking status and provide cessation support as part of sleep disorder management. Public health campaigns should include messages about the negative impact of smoking on sleep.

## Data Availability

All data utilized in this study are derived from the National Health and Nutrition Examination Survey (NHANES), which is publicly available through the Centers for Disease Control and Prevention (CDC) website. These data can be accessed freely by the public, and further details on how to access and utilize NHANES data can be found at [insert the specific URL for the NHANES data access page]. No additional unpublished data were generated during the course of this study.

https://www.cdc.gov/nchs/nhanes/index.htm

